# Somatic mutation profiles of muscle-invasive bladder cancer in Uganda: A cross-sectional study utilising whole-exome sequencing

**DOI:** 10.1101/2025.09.15.25335787

**Authors:** Badru Ssekitooleko, Moses Galukande, Hawa Nalwoga, Sam Kalungi, Noah Kiwanuka, Dan Namuguzi, Isaac Kajja, Bashir Ssuna, Frank Asiimwe, Job Kuteesa, Joseph Benon Masaba, Haruna Muwonge

## Abstract

**Background:** Muscle-invasive bladder cancer (MIBC) is a significant cause of cancer-related mortality in sub-Saharan Africa, yet its genomic landscape remains poorly characterised. In Uganda, nearly 60% of urothelial carcinoma cases are muscle-invasive (MIUC), with a near-equal sex distribution, in contrast to global trends. We aimed to define the somatic mutational landscape of Ugandan MIUC using whole-exome sequencing (WES).

**Methods:** In this descriptive cross-sectional study, 57 formalin-fixed paraffin-embedded MIUC tissue blocks were consecutively selected. DNA extraction and exome library preparation were performed for all; 47 passed quality control and 33 remained in the final dataset. Sequencing was performed on the Illumina NovaSeq 6000 platform. Reads were aligned to hg19/GRCh37, and variants called using GATK Mutect2 in tumor-only mode with stringent artefact filters. Variants were annotated with COSMIC, ClinVar, REVEL, and classified according to ACMG/AMP guidelines. Eighteen samples were resequenced on the Aviti platform to validate the low frequency of TP53 mutations. Clinical and histopathological data were also reviewed.

**Results:** All patients presented with gross hematuria, with a median age of 56 years; 17/33 (51.5%) were male. Histological variants occurred in 23/33 (69.7%), most commonly the squamous, microcystic, and nested types (each accounting for 5/23; 21.7%). LVI was present in 28/33 (84.8%). A total of 305 coding mutations were identified, averaging 9.2 ± 6.5 per tumor. Squamous and microcystic variants showed the highest burdens (16.4% and 14.4%). At the nucleotide level, 200/331 (60.4%) substitutions were C>T or C>G, consistent with APOBEC mutagenesis. Missense variants predominated (274/305; 89.8%), followed by frameshifts (31/305; 10.2%). PDE4DIP was the most frequent alteration (8/33; 24.2%), although it was not a canonical driver. Canonical recurrent drivers included FGFR3 (18.2%), FOXA1 (15.2%), BAP1 (15.2%), KMT2C (15.2%), and ERCC2 (12.1%). TP53 mutations were rare (2/33; 6.1%), and Aviti resequencing confirmed the same two variants, with additional low-confidence calls excluded. Mutations were broadly distributed, enriched on chromosomes 1, 16, 7, 19, and 17.

**Conclusion:** Ugandan MIUC displays a heterogeneous mutational profile, marked by frequent missense mutations, a high VUS burden, low TP53 frequency, and APOBEC signatures, with PDE4DIP a non-canonical gene emerging as the most frequently mutated. These findings underscore the need for African-specific genomic references to guide precision oncology.

## Introduction

Bladder cancer (BC) remains a pressing global health issue, with over 570,000 new cases and 210,000 deaths reported annually(1). Among its subtypes, muscle-invasive Urothelial Carcinoma (MIUC) represents a more aggressive form, accounting for approximately 25– 30% of all bladder cancer cases among the white population at diagnosis(2). However, for unknown reasons, higher rates (58.9%) are observed in Uganda(3). MIUC is associated with significantly worse prognosis, higher treatment costs, and increased mortality compared to the non-muscle-invasive type (4–6). Immunotherapy promises better patient outcomes in countries that can afford it; however, radical cystectomy and cisplatin-based chemotherapy remain the mainstay treatments. (5,7).

In the era of precision oncology, genomic profiling, particularly through whole-exome sequencing (WES), has become integral to cancer care by enabling the detection of somatic mutations, driver genes, and clinically actionable biomarkers, thereby facilitating personalised treatment strategies(8–10). WES has revealed recurrent alterations in genes such as TP53, FGFR3, KDM6A, ERBB2, and PIK3CA, underscoring the molecular heterogeneity of MIUC and the opportunity for personalized interventions(11,12).

While limited African genomic studies have identified unique mutational patterns such as higher frequencies of TP53, RB1, and HRAS mutations and lower FGFR3 alterations, Uganda lacks comprehensive genomic data on MIUC (7,13–15). The existing knowledge gap significantly undermines the nation’s capacity to harness advancements in precision medicine and hampers regional participation in global therapeutic trials. Furthermore, previously in Uganda, 54.3% of bladder cancer cases were identified as squamous cell carcinoma (SCC), followed by adenocarcinoma at 18.8%, urothelial carcinoma at 13.8%, and other categories comprising 13.0%(16). Current histopathological evaluations in Uganda indicate that U.C has become 11 times more prevalent than both SCC and adenocarcinoma(15). This shift highlights the need to investigate bladder cancer at the molecular level; moreover, given the high burden of muscle-invasive bladder cancer in our setting and the paucity of genomic data from African populations, this study aimed to characterise the somatic mutational landscape of Ugandan MIUC using whole-exome sequencing and to compare these patterns with global datasets.

## Methods

A descriptive cross-sectional study was conducted at the Mulago National Referral Hospital (MNRH) complex. We consecutively selected all the available 57 archived formalin-fixed paraffin-embedded (FFPE) tissue blocks of confirmed muscle-invasive urothelial carcinoma from the biorepository of MNRH, 2019-2022. Only FFPES of patients aged 18 years and above were selected. Damaged tissue blocks, those with extensive necrosis, and those with missing vital demographic data were excluded. Two labs were utilised for the study. The Mulago National Referral Hospital conducted the histopathology studies, while all whole-exome sequencing (WES) analyses were performed by Unipath Speciality Laboratory Ltd. in Ahmedabad, Gujarat, India.

The histopathologic staging was performed according to the 2004 World Health Organisation (WHO) classification(17). The retrieved FFPE tissue blocks were trimmed and cut into 4-micron-thick sections on a microtome machine. The sections were spread on the surface of water (5-10°c) below the melting point of wax to remove wrinkles, then mounted onto labelled, salinised glass slides. They were fixed by dry heat from an oven at 55-65 °C for 30-60 minutes to melt the wax, followed by dewaxing in xylene for about 5 minutes. The sections were then rehydrated in a series of graded alcohols, as follows: absolute 100%, 90% alcohol, and 70% alcohol for 1-3 minutes each. They were then washed in distilled water for a minimum of 30 seconds. Hematoxylin and Eosin (H&E) staining was done using standard operating procedures.

Two consultant pathologists examined the tissues independently, without full knowledge of the clinical data. In the event of disagreement, a consensus was achieved by re-examining the tissues using a multi-headed microscope. However, inter-observer agreement (e.g., using kappa statistics) was not formally calculated.

### Isolations and Quantitative analysis of DNA

DNA was extracted from 57 formalin-fixed paraffin-embedded (FFPE) tissue blocks using the Alexgen FFPE DNA extraction kit (CAT# AG-FF50), following the manufacturer’s instructions. Initial quality control (QC) of extracted DNA involved fluorometric quantification of double-stranded DNA using the Qubit® 4.0 Fluorometer (Thermo Fisher Scientific) and assessment of DNA integrity via 1% agarose gel electrophoresis. Samples were deemed to have passed initial QC if they demonstrated intact, high-molecular-weight DNA of at least 0.2 µg (equivalent to ≥10 ng/µL in a 20 µL eluate). All 57 samples met these extraction QC criteria and were forwarded for library preparation. (supplementary Table 1). **Preparation of the library:**

The exome libraries were prepared using the Twist Bioscience Human Core Exome Kit 2.0 (CAT# 104207). Briefly, 50 ng of high-quality genomic DNA per sample was used as input, as recommended by the manufacturer for FFPE-derived material. DNA was enzymatically sheared into smaller fragments, followed by a combined end-repair and A-tailing step, in which an adenine residue was added to the 3’ ends of the fragments to enable adapter ligation. Illumina-compatible adapters were then ligated to both ends of the DNA fragments. These adapters allow for barcoding, PCR amplification, and hybridisation to the flow cell for sequencing. Groups of eight samples were pooled together for exome capture using Twist Bioscience hybridisation probes, and the procedure was repeated until all libraries were prepared. A final amplification step was performed to enrich the captured exonic regions, and the libraries were evaluated for fragment distribution and quality using TapeStation electrophoresis. Samples that failed to generate at least 5 GB of sequencing data were excluded from analysis, as this threshold ensures sufficient depth for detecting somatic variants. After this filtering, 47 libraries passed QC and proceeded to sequencing (supplementary Table 2).

### Cluster Generation and Sequencing

The 47 qualified libraries were loaded onto the Illumina NovaSeq 6000 for cluster generation and sequencing using paired-end reads (2 × 150 bp). Paired-end sequencing enabled the template fragments to be sequenced in both the forward and reverse directions. The library molecules were bound to complementary adapter oligos on a paired-end flow cell. The adapters were designed to enable the selective cleavage of the forward strands after re-synthesis of the reverse strand during sequencing. The copied reverse strand was then used to sequence from the opposite end of the fragment. Subsequently, raw sequencing reads were produced for each library. Out of these, 42 libraries were successfully generated, exhibiting comprehensive mapping and coverage statistics appropriate for downstream analysis. The mean sequencing yield was approximately 14GB, resulting in an effective mean exome depth of approximately 100×, with a median panel coverage of 96.5% of target bases (Supplementary Table 3).

### Bioinformatics Analysis

The raw sequencing reads obtained subsequent to demultiplexing were initially evaluated for quality using FastQC v0.12.1. Adapter sequences and low-quality bases were trimmed using Cutadapt, followed by a secondary FastQC assessment to verify the efficacy of the trimming. The resulting high-quality reads were aligned to the hg19 human reference genome (GRCh37) (chosen for comparability with COSMIC and TCGA datasets) using a BWA-based alignment pipeline. Alignment files were produced in Sequence Alignment Map (SAM) format and subsequently converted to Binary Alignment Map (BAM) files. These files were sorted by coordinate, and PCR duplicates were identified and eliminated. Base quality score recalibration was conducted using the Genome Analysis Toolkit (GATK v4.3) following established best practices. Somatic single-nucleotide variants (SNVs) and small insertions/deletions (InDels) were called using Mutect2, with capture regions specified by the Twist 2.0 BED file. Mutect2 was run in tumor-only mode (without matched normals). Artefact suppression included FilterMutectCalls, strand-bias filters, and FFPE/OxoG error models.

### Variant annotation

Variant annotation was performed using GENEWERK’s proprietary annotation pipeline, which is based on the RefSeq gene model (release 109) and integrates multiple public databases. Disease associations were derived from ClinVar (release 20240310) and the Catalogue of Somatic Mutations in Cancer (COSMIC v95, GRCh37). Population allele frequencies were referenced against gnomAD v3.1.2 (exome and genome), ExAC v0.3, 1000 Genomes Phase 3, and the National Heart, Lung, and Blood Institute – Exome Sequencing Project (NHLBI-ESP) 6500 dataset. Variants present at >1% allele frequency in any population database were excluded. The functional impact of missense variants was assessed using REVEL scores accessed via dbNSFP v4.3a, with a threshold of >0.5 used to indicate potential pathogenicity, corresponding to a sensitivity of 0.754 and a specificity of 0.891 (18).

### Variant filtering

High-confidence somatic calls were defined as those with ≥10 paired supporting reads, a total depth of ≥50× at the variant site, balanced forward/reverse strand representation, and the absence of mapping or sequencing artefact flags. All variants were filtered through a curated somatic cancer gene panel and classified in accordance with consensus guidelines from the American College of Medical Genetics and Genomics (ACMG) and the Association for Molecular Pathology (AMP)(19,20). Only variants classified as pathogenic, likely pathogenic, or variants of uncertain significance (VUS) with phenotypic relevance were retained for downstream interpretation. Final variant reporting was conducted at three annotation levels: gene symbol (HGNC), transcript/cDNA (HGVS c. notation), and protein (HGVS p. notation), enabling functional interpretation, cross-study comparability, and alignment with international reporting standards. Initial variant calling across the 42 libraries produced variable numbers of candidate variants per sample. PASS calls ranged widely, reflecting heterogeneity in DNA quality and tumor content. These intermediate variant counts are summarised in Supplementary Table 4. Following application of high-confidence somatic criteria, nine libraries were excluded for low callable regions or insufficient depth, leaving 33 libraries for final downstream analysis.

All 57 FFPE tissue blocks from patients with muscle-invasive bladder cancer successfully passed DNA quality control. Among these, 47 libraries met the criteria for library preparation quality control (≥5 Gb threshold). Subsequently, 42 libraries produced comprehensive mapping and coverage statistics, and 33 libraries yielded high-confidence somatic variant calls appropriate for downstream analysis.

### Statistical Analysis

This study employed a descriptive framework aimed at characterising the somatic mutational landscape of muscle-invasive bladder cancer (MIUC) in a Ugandan cohort using whole-exome sequencing. Variant counts, frequencies, chromosomal distributions, and functional classifications were summarised using descriptive statistics. References to external datasets, such as the TCGA, were used for exploratory and descriptive purposes.

### Cross-Platform Validation and Analytical Quality Control

Given the unexpectedly low frequency of TP53 mutations observed in the initial Illumina-based Whole-Exome Sequencing (WES) of 33 samples, a subset of 18 samples was randomly selected for resequencing on the Aviti platform to evaluate potential platform-specific biases (supplementary Table 5). The Aviti system consistently detected both TP53 mutations identified by Illumina. Additionally, Aviti identified three further TP53 variants; however, none met our pre-specified high-confidence criteria (≥10 paired supporting reads, total depth ≥50×, balanced strand representation, absence of artefact flags) and were therefore excluded from downstream analysis. The majority of these additional findings were low-support variants (1–6 paired reads; depths 9–36×) located in coding, intronic, promoter, or untranslated regions (UTRs)

Furthermore, across the broader gene set, Aviti identified a larger spectrum of somatic mutations in other muscle-invasive bladder cancer–related genes, which likely reflects its deeper coverage, improved library preparation, and better handling of FFPE-derived DNA. This cross-platform validation confirmed that high-confidence variant calls were reproducible between platforms and that the absence of additional TP53 mutations was not due to platform-specific under-detection.

Sequencing and variant interpretation were conducted under the supervision of two senior consultants in clinical genetics and molecular biology (Unipath Speciality Laboratory Ltd., Ahmedabad, India).

## Results

### Clinicopathologic Characteristics of Study Participants

A total of 33 formalin-fixed paraffin-embedded (FFPE) muscle-invasive bladder cancer (MIBC) tumor samples were included in the final analysis (Figure 1). The median age of patients was 56 years (range: 30–98), with a nearly equal gender distribution (17 males, 16 females). All patients presented with haematuria and a clinically or radiologically detectable bladder mass. On clinical and pathologic assessment, all tumors were staged as T2 or higher, consistent with the inclusion criteria for MIUC. Most samples exhibited histologic variant differentiation, with 23/33 (69.7%) cases, while only 10 cases (30.3%) were classified as pure urothelial carcinoma. The most frequently observed variants included squamous differentiation, microcystic pattern, and nested variant, each accounting for 5/23(21.7%). Lymphovascular invasion (LVI) was identified in 28 of 33 cases (84.8%). (Supplementary Table 6)

**Figure 1:**
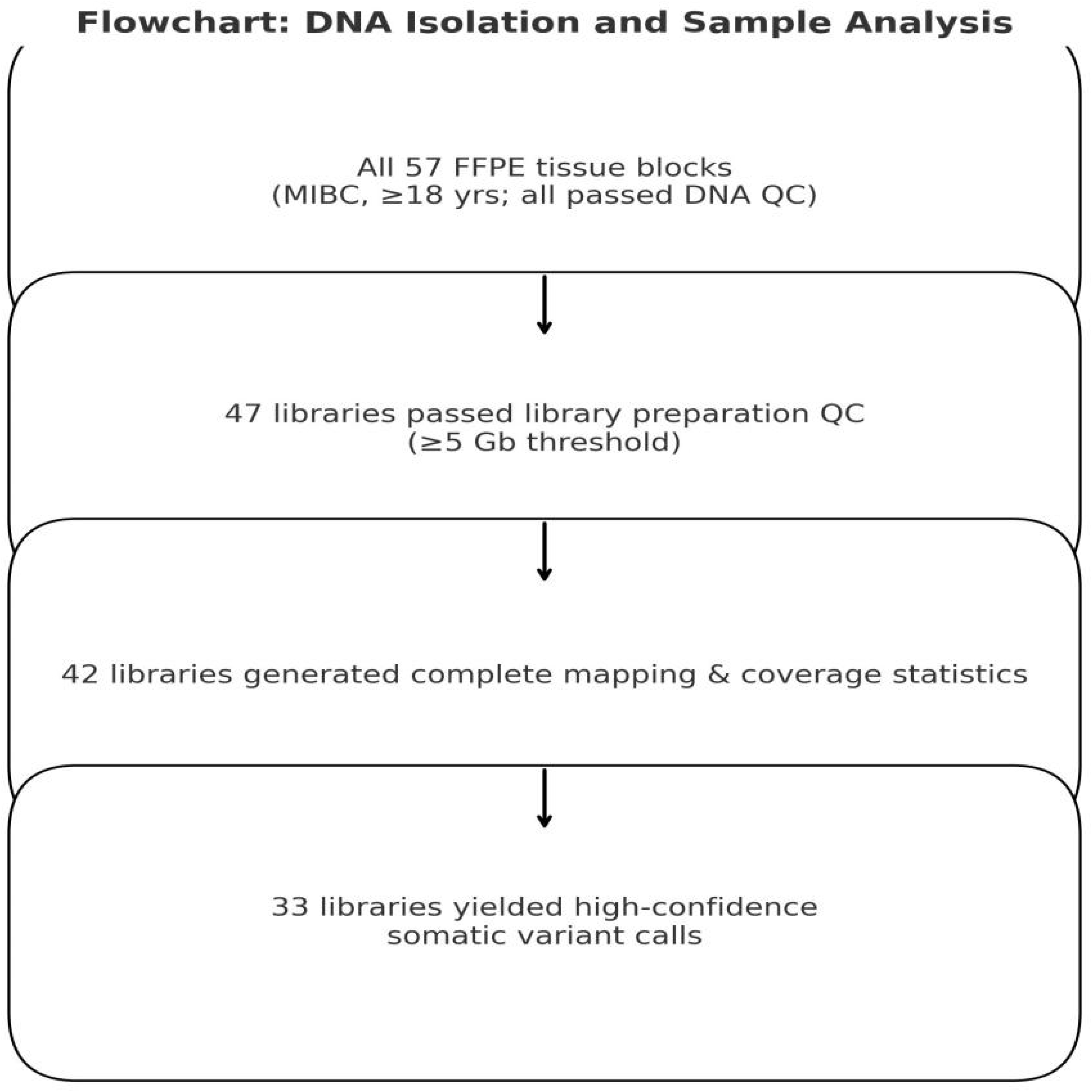
Flowchart of DNA isolation and sequencing analysis.

### Somatic Tumor Mutation Burden

Across the cohort, a total of 305 somatic coding mutations were identified, with an average mutation count of 9.2 ± 6.6 mutations per sample. Mean mutation counts were generally similar between tumors with histologic variant differentiation and pure urothelial carcinomas (9.4 ± 6.4 vs. 8.8 ± 7.3 mutations per sample, respectively). Squamous differentiation and microcystic variants exhibited the highest mutation burden among the histologic variants at 50/305 (16.4%) and 44/305 (14.4%), respectively.

Strand-normalised nucleotide substitutions were dominated by transitions (C>T and T>C), which accounted for 200/331(60.4%) mutations, while transversions (C>A, C>G, T>A, T>G) comprised 131/331 (39.6%). Frameshift mutations were the most frequent type of indel, accounting for 31/305 (10.2%). Missense mutations predominated among amino acid changes (84.3%), and the majority of variants were classified as variants of uncertain significance (VUS; 93.8%). The 10 (3.3%) pathogenic variants include TP53, STAG2, ATM, FGFR3, RUNX1, PIK3CA, FOXA1, ERBB2, ASXL1, and HRAS. The 9 (3.0%) likely pathogenic variants include KMT2C, KMT2D, BAP1, ARID1A, FANCA, RB1, PDE4DIP, FGFR2, and SETBP1. The remainder are VUS. (Supplementary Table 7)

The genomic landscape revealed variants distributed across nearly all chromosomes, with distinct patterns of enrichment and dispersion. Notably, chromosomes 1 (37 mutations; 12.1%), 16 (25; 8.2%), 7 (24; 7.9%), 19 (23; 7.5%), and 17 (23; 7.5%) exhibited the highest mutation frequencies, collectively accounting for approximately 43% of all identified variants. Conversely, chromosomes 20 (5; 1.6%), 5 (5; 1.6%), and 21 (3; 1.0%) exhibited the fewest mutations.

The bar chart shows mutation frequencies for the 24 most frequently altered genes, grouped by chromosomal location. The most commonly altered genes were PDE4DIP (8 events, chr1), FGFR3 (6 events, chr4), FOXA1 (5 events, chr14), TYRO3 (5 events, chr15), BAP1 (5 events, chr3), KMT2C (5 events, chr7), and MKI67 (5 events, chr10). Additional recurrent alterations (frequency 3–4) were observed in ERCC2, SEPTIN9, CUX1, ARID1A, TERT, and others. Mutations were dispersed across multiple chromosomes, with higher representation on chromosomes 1, 3, 4, 7, 10, 14, and 15. (Supplementary Table 7).

## DISCUSSION

The study investigated the genetic landscape of muscle-invasive bladder cancer (MIUC) in Uganda through whole-exome sequencing (WES). Our findings expand the limited genomic data available from African populations and highlight both shared and unique features compared to global cohorts.

The average mutation count per sample was 9.2 ± 6.6, which is within the range of other previous WES-based bladder cancer studies(10,21). Notably, variants classified as of uncertain significance (VUS) constituted the majority (93.8%), consistent with findings from underrepresented populations, where existing variant databases have limited representation of African genomes(8,14). This highlights the need for expanded African genomic databases and reference cohorts to enhance the accuracy of variant interpretation.

### Mutational Burden and Histologic Variants

Squamous differentiation and microcystic variants demonstrated the highest mutation burdens, accounting for 16.4% and 14.4% of all identified mutations, respectively. These findings support earlier studies showing that variant histologies in MIBC are often associated with higher genomic instability and worse clinical outcomes(5,22). Lymphovascular invasion (LVI), which was present in nearly 88% of cases in this cohort, underscores the aggressive phenotype of the disease and has been shown in other studies to correlate with metastatic potential and reduced survival(9).

### Pathogenic and Likely Pathogenic Variants

The ten pathogenic mutations identified included, among others, TP53, FGFR3, ERBB2, and STAG2 (Figure 1C and Supplementary Table 8). Unexpectedly, TP53, despite being one of the most frequently mutated genes in global MIBC datasets, was found to be mutated in only 2 of 33 samples (6%). This striking underrepresentation contrasts sharply with The Cancer Genome Atlas (TCGA) and other large international cohorts, in which TP53 mutations are observed in 49–60% of cases(10,12). The low mutation frequency observed here aligns with our earlier findings from TP53 immunohistochemistry (IHC), where protein overexpression did not correspond to genomic alterations(7). This discrepancy may be explained by wild-type TP53 stabilisation, β-catenin signalling, or epigenetic modifications(23,24).

### Somatic Mutation Patterns and Indels

Missense (non-synonymous) mutations represented the most prevalent amino acid–level alterations, constituting 84.3% of the coding mutations (85.1%) (Figure 1B). These protein-altering variants are believed to confer a selective advantage in tumor evolution by disrupting pathways associated with cell cycle control, DNA repair, and growth signalling patterns that are consistent with previous analyses conducted by The Cancer Genome Atlas (TCGA) and Asian bladder cancer cohorts(10,21,25). Frameshift mutations were identified as the most frequent type of insertion/deletion (indel) events, accounting for 31/305 (10.2%) mutations. These alterations typically result in truncated, non-functional proteins and are frequently associated with the inactivation of tumor suppressor genes, aggressive tumor phenotypes, and poor prognosis(26). Additional mutation types encompassed synonymous mutations (11/305; 3.6%), stop-gain mutations (5/305; 1.6%), and in-frame indels (0.3%), each contributing minimally to the overall mutational landscape. These observations emphasise the predominance of protein-altering mutations in MIBC evolution and may inform future precision oncology strategies targeting specific mutational profiles.

### Evidence of APOBEC Mutagenesis

Although this study did not perform formal mutational signature decomposition based on trinucleotide context, the observed substitution spectrum (Figure 1A) is consistent with established APOBEC-associated signatures (COSMIC SBS2 and SBS13). In particular, C>T (52.9%) and C>G (7.9%) substitutions are hallmark features of APOBEC cytidine deaminase activity. This indicates a substantial contribution of APOBEC-driven mutagenesis in this Ugandan MIUC cohort. However, it remains lower than the approximately 80% APOBEC-related burden reported in TCGA muscle-invasive bladder cancer datasets (27). This mutational process has previously been associated with higher tumor mutational burden, immune infiltration, and potential responsiveness to immunotherapy(28,29).

### Classification of Mutated Genes by Biological Function and Frequency

When grouped by biological function, some pathways were only modestly represented in our dataset, which contrasts with the findings of many cohorts (Supplementary Table 7).

- **Apoptosis and DNA damage response:** Genes like TP53, ATM, BAP1, and FANCA were rarely mutated in this cohort. For instance, TP53 appeared in only two samples (6%), in contrast to its near-universal alteration in global datasets. This suggests that Ugandan tumors may rely less on canonical apoptotic escape mechanisms and more on alternative pathways for survival and progression.
- **Chromatin remodelling and epigenetic regulation:** Our dataset identified mutations in a limited number of genes related to chromatin remodelling and epigenetic regulation, such as KMT2C, KDM6A, KMT2D, ARID1A, and STAG2. These genes encompass essential functional categories such as histone modifiers (KMT2C, KMT2D, KDM6A), components of the SWI/SNF complex (ARID1A), and cohesin complex genes (STAG2). The overall mutation rate remained low, indicating minimal disruption of the usual epigenetic pathways in this group. This stands in contrast to larger genomic studies like TCGA, which report recurrent mutations in chromatin modifier genes in 25–35% of bladder tumors (10,12). Notably absent from our dataset are canonical chromatin regulators frequently observed in other bladder cancer studies, including EZH2, SMARCA4, CHD6, CHD7, and SMC1A(10,21). The absence of these mutations and the low mutation rate of the genes listed above, particularly those linked to aggressive tumor behaviour and immune modulation, may indicate population-specific epigenetic profiles underscoring the necessity for broader genomic and epigenomic research that includes African populations.
- **Signal transduction and growth regulation:** FGFR3 mutations were detected in **18.2%** of our Ugandan MIBC cases, aligning closely with the 21% frequency reported by TCGA, particularly in the luminal papillary subtype. Though typically linked to non-muscle invasive bladder cancer (NMIBC), FGFR3 alterations in MIBC suggest a broader role in bladder cancer progression and highlight potential benefit from FGFR-targeted therapies such as erdafitinib(10). Other signal transduction genes such as PIK3CA, ERBB2, HRAS, EGFR, BRAF, ERBB3, NF1, and PTEN were also present, although mutated at lower frequencies. In contrast, well-known signal transduction regulators often reported in global MIBC studies, such as KRAS, NRAS, AKT1, and RAF1, were notably absent in this cohort(10,27). This may imply a more limited role for these pathways in the progression of MIBC in Ugandan patients, which could reduce the utility of specific targeted therapies that are otherwise more commonly applicable in Western populations(10).
- **Cell cycle regulation:** Our dataset revealed mutations in several genes involved in cell cycle control, including RB1, CDKN1A, CDKN2A, TP53, CCND1, and ATM. However, the overall mutation frequency of these genes was low, with several samples lacking any detectable alterations in cell cycle regulatory genes. These genes play crucial roles in regulating the G1/S checkpoint and maintaining genomic integrity. However, other key regulators that are frequently mutated in global MIBC studies, such as E2F3, MDM2, CDK4, CDK6, CDK2, and CDKN2B, were absent in our cohort (10–12). The absence of some of these cell cycle genes suggests possible reliance on alternative regulatory mechanisms or involvement of non-coding and epigenetic alterations, which may define a region-specific pathogenesis of MIBC in Ugandan patients.

It is important to emphasise that although some genes are functionally significant in the context of bladder cancer biology, their actual mutation frequency in this Ugandan cohort was limited. Therefore, caution is warranted in extrapolating biological importance solely from Western datasets to African populations.

### Genomic distribution across Chromosomes

Somatic mutations were distributed across nearly all chromosomes (Figure 2). Notably, Chromosome 1 exhibited the highest mutation frequency (37/305; 12.1%), followed closely by Chromosomes 16 and 7 (8.2% and 7.9%, respectively). This widespread distribution suggests that the mutational processes driving MIUC in this population are not confined to specific genomic loci but are instead broadly dispersed throughout the genome. Such chromosomal spread is consistent with findings from previous genomic studies of bladder cancer. Deletions on Chromosome 9, particularly at 9p21 involving CDKN2A and DBC1, are among the earliest and most frequent events in urothelial carcinoma, occurring in up to 60% of tumors (30,31). Additional chromosomal imbalances involving Chromosomes 3, 7, 13, and 17 have also been associated with tumor progression and recurrence, suggesting their role in disease aggressiveness(32,33). Moreover, the comprehensive TCGA analysis confirmed that bladder cancer exhibits substantial chromosomal instability, including widespread copy number alterations and aneuploidy across the genome.(27) . These alterations collectively contribute to tumor heterogeneity, reinforcing the importance of evaluating chromosomal alterations when assessing tumor evolution and therapeutic opportunities, especially within underrepresented populations.

**Figure 2:**
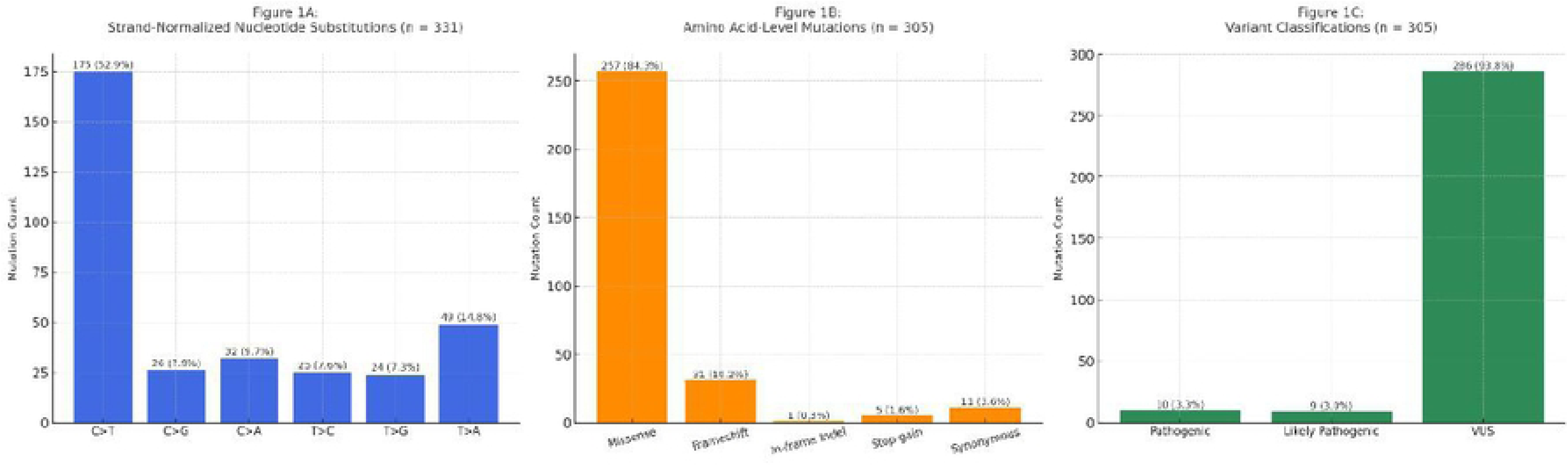
Distribution of Nucleotide Mutations, Amino Acid Mutations, and Genetic Variants.

**Figure 3:**
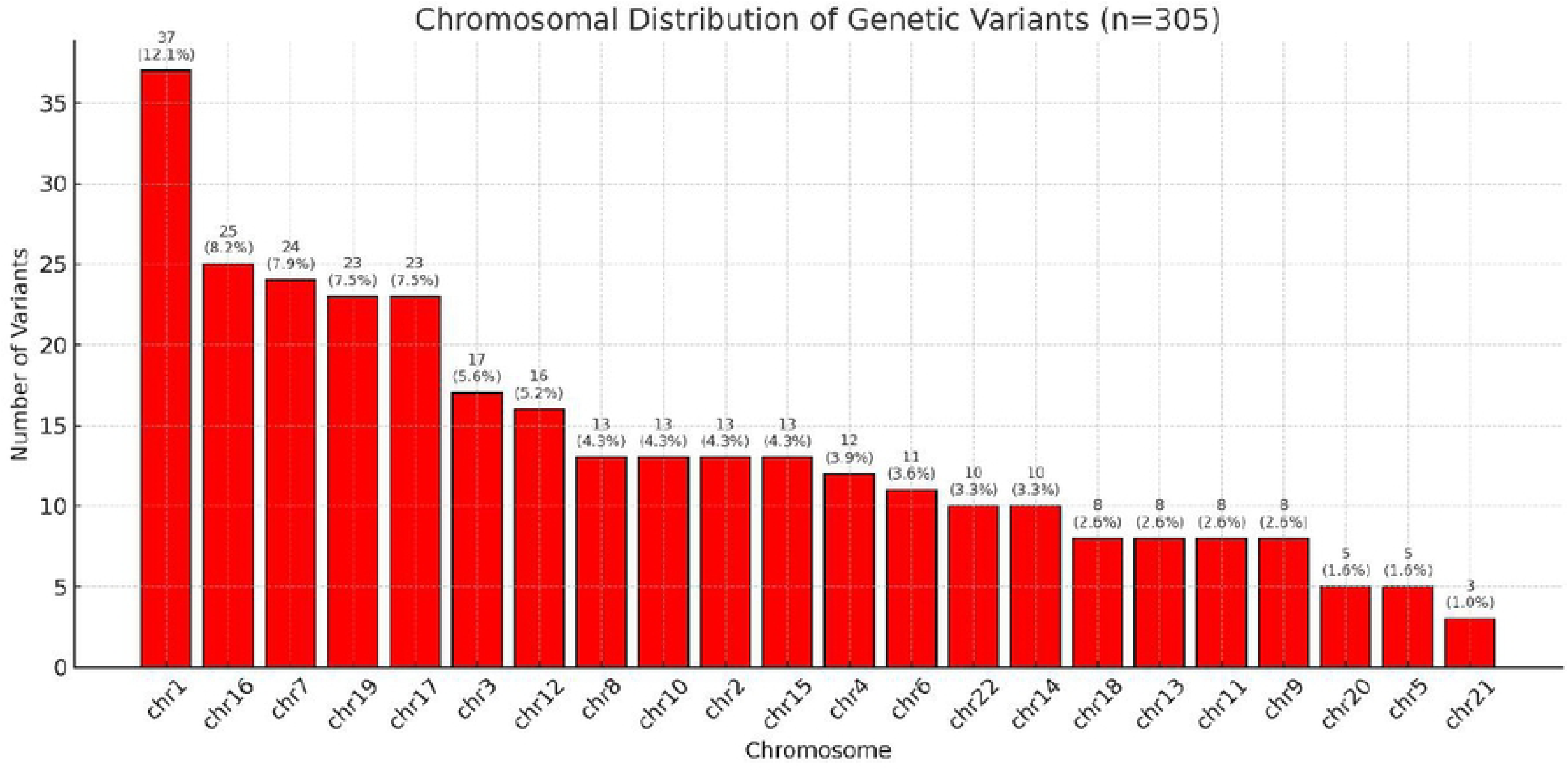
Chromosomal distribution of somatic variants.

**Figure 4:**
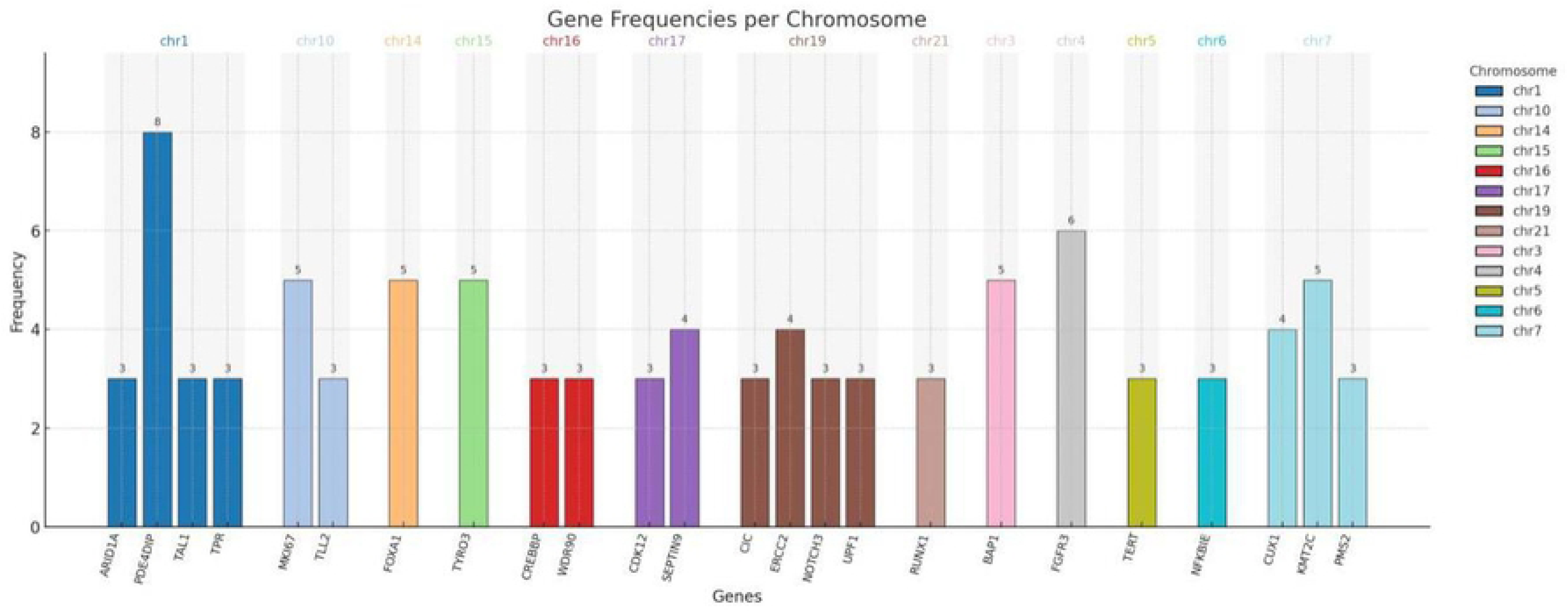
Recurrently mutated genes by chromosomal location.

### Limitations

Our study, albeit groundbreaking in the region, has a modest sample size that may limit the generalisability of our findings. Although intratumoral heterogeneity can influence mutational profiles, we extracted DNA from more than two representative FFPE blocks per tumor, thereby reducing the likelihood of missing subclonal variants. The absence of transcriptomic data limited direct validation of APOBEC mutagenesis and gene expression signatures, though mutational context analyses provided indirect support. Furthermore, the lack of matched normal tissue or a panel of normals necessitated tumor-only variant calling, which may have inflated the number of VUS. Their interpretation was also constrained by the limited representation of African genomes in major reference databases; nonetheless, we applied stringent annotation filters and cross-referenced multiple sources to mitigate misclassification.

## Conclusion

This study reveals a heterogeneous somatic mutational landscape in Ugandan MIBC, characterised by a predominance of missense mutations, a high burden of variants of uncertain significance, and an unusually low frequency of TP53 alterations compared with global datasets. Substitution patterns suggest APOBEC-associated mutagenesis, underscoring the role of population-specific mutational processes. Notably, PDE4DIP, a non-canonical driver of MIUC, was the most frequently mutated gene in this cohort, suggesting potential regional or context-specific biology that warrants further investigation. Clinically actionable alterations, including FGFR3 and ERBB2, were also identified. Together, these findings highlight distinct features of Ugandan MIUC and provide both novel biological insights and potential therapeutic entry points for precision oncology in Africa.

## Data Availability

All relevant data are within the manuscript and its Supporting Information files.

## Declarations

**Ethical clearance**: Clearance was obtained from Makerere University School of Medicine research and ethics committee under protocol reference number **Mak-SOMREC-2021-257**

## Plagiarism

The authors confirm that the work submitted is original and does not violate the journal’s plagiarism policy.

## Authors contributions

**Principle investigator**: Badru Ssekitooleko (BS)
**Development of the study protocol:** BS, bs, NK, HM, HN, MG, IK, FA, SK
**Data collection:** BS, JK, JBM, FA, DN, SK
**Data analysis:** bs, BS, NK.
**Manuscript writing**: all authors contributed.

## Acknowledgements

Urology fellows (Drs. Medeyi Vicent, Namugga Martha, Nassaka Victo, Tinka Ann, and all nurses working on the urology ward of Mulago National Referral Hospital). A special thank you to the staff of the pathology department.

## REFERENCES

1. Sung H, Ferlay J, Siegel RL, Laversanne M, Soerjomataram I, Jemal A, et al. Global Cancer Statistics 2020: GLOBOCAN Estimates of Incidence and Mortality Worldwide for 36 Cancers in 185 Countries. CA Cancer J Clin. 2021 May;71(3):209–49.

2. Cassell A, Yunusa B, Jalloh M, Mbodji MM, Diallo A, Ndoye M, et al. Non-Muscle Invasive Bladder Cancer: A Review of the Current Trend in Africa. World J Oncol. 2019 June;10(3):123–31.

3. Ssekitooleko B, Ssuna B, Nimanya SA, Kiwewa R, Ssewanyana Y, Nkonge E, et al. High incidence of acute kidney injury among patients with major trauma at Mulago National Referral Hospital, Uganda: risk factors and overall survival. African Health Sciences. 2022 Dec 22;4(4):191–8.

4. Aggen DH, Drake CG. Biomarkers for immunotherapy in bladder cancer: a moving target. Journal for ImmunoTherapy of Cancer. 2017 Nov 21;5(1):94.

5. Kamat AM, Hahn NM, Efstathiou JA, Lerner SP, Malmström PU, Choi W, et al. Bladder cancer. Lancet. 2016 Dec 3;388(10061):2796–810.

6. Rubino S, Kim Y, Zhou J, Dhilon J, Li R, Spiess P, et al. Positive Ki-67 and PD-L1 expression in post-neoadjuvant chemotherapy muscle-invasive bladder cancer is associated with shorter overall survival: a retrospective study. World J Urol. 2021 May;39(5):1539–47.

7. Ssekitooleko B, Nalwoga H, Kalungi S, Kiwanuka N, Galukande M, Namuguzi D, et al. Unravelling bladder cancer in Uganda: insights from the discrepancies in TP53 assessment between immunohistochemistry and whole exome sequencing. African Urology. 2025 Feb 20;5(1):5–9.

8. Abubakar SD, Bello ZM, Gusau SS, Kabir IM. Exploring racial disparities in bladder urothelial cancer: insights into survival and genetic variations. Afr J Urol. 2024 May 21;30(1):28.

9. Cheng L, Davison DD, Adams J, Lopez-Beltran A, Wang L, Montironi R, et al. Biomarkers in bladder cancer: translational and clinical implications. Crit Rev Oncol Hematol. 2014 Jan;89(1):73–111.

10. Robertson AG, Kim J, Al-Ahmadie H, Bellmunt J, Guo G, Cherniack AD, et al. Comprehensive Molecular Characterization of Muscle-Invasive Bladder Cancer. Cell. 2017 Oct 19;171(3):540–556.e25.

11. Rodriguez-Vida A, Lerner SP, Bellmunt J. The Cancer Genome Atlas Project in Bladder Cancer. Cancer Treat Res. 2018;175:259–71.

12. Weinstein JN, Akbani R, Broom BM, Wang W, Verhaak RGW, McConkey D, et al. Comprehensive molecular characterization of urothelial bladder carcinoma. Nature. 2014 Mar;507(7492):315–22.

13. Bowa K, Kachimba JS, Labib MA, Mudenda V, Chikwenya M. The pattern of urological cancers in Zambia. Afr J Urol. 2009 June;15(2):84–7.

14. Rotimi SO, Rotimi OA, Salhia B. A Review of Cancer Genetics and Genomics Studies in Africa. Front Oncol. 2021 Feb 15;10:606400.

15. Ssekitooleko B, Namuguzi D, Kalungi S, Kiwanuka N, Muwonge H, Galukande M, et al. A histopathological study of bladder cancer in Uganda. African Urology. 2024 Mar 26;4(1):1–6.

16. Anthony PP. Malignant Tumours of the Kidney, Bladder and Urethra. In: Templeton AC, editor. Tumours in a Tropical Country: A Survey of Uganda 1964–1968 [Internet]. Berlin, Heidelberg: Springer; 1973 [cited 2022 Nov 20]. p. 145–70. (Recent Results in Cancer Research). Available from: 10.1007/978-3-642-80725-1_8

17. Lopez-Beltran A, Luque RJ, Alvarez-Kindelan J, Quintero A, Merlo F, Requena MJ, et al. Prognostic factors in survival of patients with stage Ta and T1 bladder urothelial tumors: the role of G1-S modulators (p53, p21Waf1, p27Kip1, cyclin D1, and cyclin D3), proliferation index, and clinicopathologic parameters. Am J Clin Pathol. 2004 Sept;122(3):444–52.

18. Ioannidis NM, Rothstein JH, Pejaver V, Middha S, McDonnell SK, Baheti S, et al. REVEL: An Ensemble Method for Predicting the Pathogenicity of Rare Missense Variants. The American Journal of Human Genetics. 2016 Oct;99(4):877–85.

19. Li MM, Datto M, Duncavage EJ, Kulkarni S, Lindeman NI, Roy S, et al. Standards and Guidelines for the Interpretation and Reporting of Sequence Variants in Cancer: A Joint Consensus Recommendation of the Association for Molecular Pathology, American Society of Clinical Oncology, and College of American Pathologists. J Mol Diagn. 2017 Jan;19(1):4–23.

20. Richards S, Aziz N, Bale S, Bick D, Das S, Gastier-Foster J, et al. Standards and guidelines for the interpretation of sequence variants: a joint consensus recommendation of the American College of Medical Genetics and Genomics and the Association for Molecular Pathology. Genet Med. 2015 May;17(5):405–24.

21. Guo G, Sun X, Chen C, Wu S, Huang P, Li Z, et al. Whole-genome and whole-exome sequencing of bladder cancer identifies frequent alterations in genes involved in sister chromatid cohesion and segregation. Nat Genet. 2013 Dec;45(12):1459–63.

22. Choi W, Porten S, Kim S, Willis D, Plimack ER, Hoffman-Censits J, et al. Identification of Distinct Basal and Luminal Subtypes of Muscle-Invasive Bladder Cancer with Different Sensitivities to Frontline Chemotherapy. Cancer Cell. 2014 Feb 10;25(2):152– 65.

23. Bouchalova P, Nenutil R, Muller P, Hrstka R, Appleyard MV, Murray K, et al. Mutant p53 accumulation in human breast cancer is not an intrinsic property or dependent on structural or functional disruption but is regulated by exogenous stress and receptor status. J Pathol. 2014 July;233(3):238–46.

24. Xue Y, San Luis B, Lane DP. Intratumour heterogeneity of p53 expression; causes and consequences. The Journal of Pathology. 2019;249(3):274–85.

25. Xu N, Yao Z, Shang G, Ye D, Wang H, Zhang H, et al. Integrated proteogenomic characterization of urothelial carcinoma of the bladder. J Hematol Oncol. 2022 June 3;15(1):76.

26. Martincorena I, Campbell PJ. Somatic mutation in cancer and normal cells. Science. 2015 Sept 25;349(6255):1483–9.

27. Cancer Genome Atlas Research Network. Comprehensive molecular characterization of urothelial bladder carcinoma. Nature. 2014 Mar 20;507(7492):315–22.

28. Glaser AP, Fantini D, Wang Y, Yu Y, Rimar KJ, Podojil JR, et al. APOBEC-mediated mutagenesis in urothelial carcinoma is associated with improved survival, mutations in DNA damage response genes, and immune response. Oncotarget. 2017 Dec 16;9(4):4537–48.

29. Roberts SA, Lawrence MS, Klimczak LJ, Grimm SA, Fargo D, Stojanov P, et al. An APOBEC cytidine deaminase mutagenesis pattern is widespread in human cancers. Nat Genet. 2013 Sept;45(9):970–6.

30. Knowles MA. Molecular subtypes of bladder cancer: Jekyll and Hyde or chalk and cheese? Carcinogenesis. 2006 Mar;27(3):361–73.

31. Veltman JA, Fridlyand J, Pejavar S, Olshen AB, Korkola JE, DeVries S, et al. Array-based comparative genomic hybridization for genome-wide screening of DNA copy number in bladder tumors. Cancer Res. 2003 June 1;63(11):2872–80.

32. Li Y, Sun L, Guo X, Mo N, Zhang J, Li C. Frontiers in Bladder Cancer Genomic Research. Front Oncol [Internet]. 2021 May 20 [cited 2025 May 5];11. Available from: https://www.frontiersin.orghttps://www.frontiersin.org/journals/oncology/articles/10.3389/fonc.2021.670729/full

33. Linn JF, Sesterhenn I, Mostofi FK, Schoenberg M. The molecular characteristics of bladder cancer in young patients. J Urol. 1998 May;159(5):1493–6.

